# Leveraging Reddit data for Context-enhanced Synthetic Health Data Generation to Identify Low Self Esteem

**DOI:** 10.1101/2025.10.15.25338111

**Authors:** Muskan Garg, Xingyi Liu, Eunji Jeon, Joanna M. Biernacka, Mark A. Frye, Yonas E. Geda, Sunghwan Sohn

## Abstract

Low self-esteem (LoST) is a latent yet critical psychosocial risk factor that predisposes individuals to depressive disorders. Although structured tools exist to assess self-esteem, their limited clinical adoption suggests that relevant indicators of LoST remain buried within unstructured clinical narratives. The scarcity of annotated clinical notes impedes the development of natural language processing (NLP) models for its detection. Manual chart reviews are labor-intensive and large language model (LLM)-driven (weak) labeling raises privacy concerns. Past studies demonstrate that NLP models trained on LLM-generated synthetic clinical notes achieve performance comparable to, and sometimes better than those trained on real notes. This highlights synthetic data’s utility for augmenting scarce clinical corpora while reducing privacy concerns. Prior efforts have leveraged social media data, such as Reddit, to identify linguistic markers of low self-esteem; however, the linguistic and contextual divergence between social media and clinical text limits the generalizability of these models. To address this gap, we present a novel framework that generates context-enhanced synthetic clinical notes from social media narratives and evaluates the utility of small language models for identifying expressions of low self-esteem. Our approach includes a mixed-method evaluation framework: (i) structure analysis, (ii) readability analysis, (iii) linguistic diversity, and (iv) contextual fidelity of LoST cues in source Reddit posts and synthetic notes. This work offers a scalable, privacy-preserving solution for synthetic data generation for early detection of psychosocial risks such as LoST and demonstrates a pathway for translating mental health signals in clinical notes into clinically actionable insights, thereby identifying patients at risk.

## 1 Introduction

Self-esteem is a psychological construct of a person’s perceived self-worth against an external “yardstick” of success and entails comparing oneself with others. While positive self-esteem may have its mental health benefits, low self-esteem makes a person vulnerable to poor mental health outcomes. A competing theory is the construct of self-compassion that generated several publications over the past two decades ever since it was first reported by Kristin Neff. The purpose of this paper is on LoST therefore; we would refer to the reader elsewhere regarding the empirically validated mindfulness based self-compassion model (Neff, 2023). Low self-esteem is a pervasive psychosocial vulnerability as epidemiological studies estimate its prevalence among general populations is ∼34–41% in diverse settings and is strongly associated with depressive symptoms across longitudinal studies (H. Gu et al., 2024; Tunçgenç et al., 2023). Major depressive disorder affected approximately 163 million people globally in 2017 (∼2 % of the world population at that time) and was identified as the second-leading contributor to years lived with disability (Yan et al., 2024). Past studies identified low self-esteem as a causal factor in depression (H. Gu et al., 2024; Lee et al., 2021; Liu et al., 2024; Tunçgenç et al., 2023). Despite its recognized importance, low self-esteem faces a lack of consensus definition. Self-esteem is measured via self-report scales like the Rosenberg Self-Esteem Scale (RSES) but they are rarely documented as structured clinical assessments in clinical practice. In routine visits, clinicians rarely administer structured instruments and often do not explicitly document self-esteem, even when relevant indicators are present and observable (de la Cámara & Lobo, 2020). As a result, semantic markers of low self-esteem are buried in unstructured clinical notes.

This limitation poses a significant challenge for hospital systems, where mental health conditions frequently co-occur with chronic disease, emergency visits, or inpatient care. Timely identification of latent psychosocial risk factors such as low self-esteem can improve patient triage, care coordination, and personalized treatment planning. Detecting these cues in existing clinical narratives would greatly enable proactive psychosocial intervention and support patient care. However, developing models to extract low self-esteem from clinical notes is fundamentally limited by the lack of annotated data. Annotating real clinical notes is labor-intensive, expensive, and subject to privacy, legal, and institutional challenges.

Prior research in clinical text extraction, such as applying natural language processing (NLP) to identify mental illness or substance use from electronic health record (EHR) notes, has demonstrated that unstructured clinical narratives contain valuable signals often missed in structured fields (Newby et al., 2024; Seinen et al., 2025; Shankar et al., 2025; X. Zhang et al., 2024). Similarly, synthetic clinical note generation from deidentified or literature-based sources has enabled model training while preserving privacy (Nadăş et al., 2025). Past studies used synthetic notes to train clinical language models with promising downstream performance on real data (Kang et al., 2024). On the other hand, social media platforms, particularly Reddit, have been widely studied for self-esteem and depression signals using NLP (Garg, 2023; Garg et al., 2023; Garg et al., 2024). These community-generated narratives offer rich linguistic markers of low self-esteem, but the narrative tone, stylistic/linguistic features, and content differ markedly from clinical notes (see Appendix A).

Consequently, models trained on Reddit data do not generalize well to clinical text domains. These differences in the nature of the text and the absence of annotated clinical notes remain major barriers. This gap in knowledge motivates our studies. To address this bottleneck, we propose curating synthetic clinical notes from publicly available Reddit posts, which are already rich in first-person narratives expressing low self-esteem and its linguistic markers. This conversion enables the development of NLP models for clinical use by mitigating the potential leak of protected health information and circumventing institutional data constraints. Also, this approach provides a scalable source of weak supervision, which can seed initial model development and evaluation for clinical notes in the absence of gold-standard clinical annotation. The significance of this synthetic data generation lies in its ability to study the linguistic and contextual divide between community-generated data and clinical documentation. We conceptualize a pipeline: (1) transform social media narratives into context-enhanced synthetic clinical notes; (2) comparative analysis of small language models in extracting LoST, deployed on Reddit posts and synthetic notes, to identify low self-esteem (see Figure 1).

**Figure 1.**
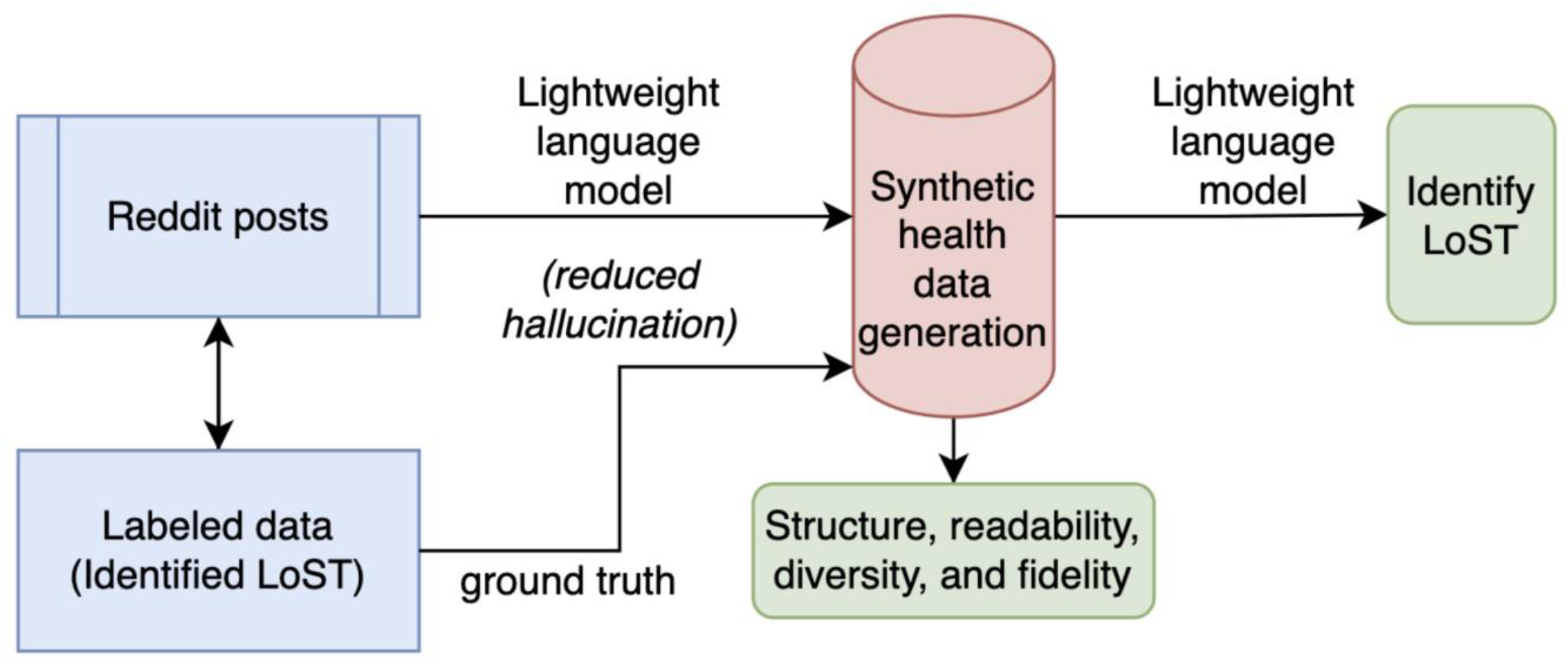
Overview of the proposed work.

This approach opens a new frontier in synthetic data–driven clinical NLP, enabling practical and efficient modeling of under-documented psychosocial indicators for real-world clinical impact. Our major contributions are as follows:

- We propose a novel framework for converting Reddit mental-health narratives into synthetic clinical notes, addressing privacy, scalability, and domain-adaptation needs.
- We assess the linguistic, structural, and semantic alignment between Reddit posts and context-enhanced synthetic clinical notes, emphasizing differences in stylometric patterns across the datasets.
- We test and validate the efficacy of the state-of-the-art small language models (SLMs) on Reddit posts and synthetic notes to extract low self-esteem indicators.

We demonstrate that our synthetic data-driven approach enables privacy-preserving, cost-ands time-efficient detection of low self-esteem in clinical text. By leveraging psychological self-assessment - rosenberg self-esteem scale (RSES) driven ground truth for synthetic notes, this study addresses a critical gap and offers (i) generation of synthetic notes, and (ii) an empirical model for early psychosocial low self-esteem detection in mental health analysis.

### Related work

Social Media Signals of Low Self-esteem: Prior work fine-tuned GPT model for suicide risk detection in social media posts, demonstrates the promise of transformer-based models while also revealing their limitations in capturing implicit signals like hopelessness (Howard et al., 2020). Recent reviews affirm that deep learning models outperform traditional approaches in social media-based mental health detection, but domain-specific adaptation remains critical for identifying psychosocial expressions (Zhang et al., 2022). Social media platforms such as Reddit have emerged as valuable sources for detecting early signs of mental health issues, including depression and related psychosocial constructs (Eichstaedt et al., 2018). The recently introduced LoST dataset provides expert-annotated Reddit posts labeled for low self-esteem using psychology-informed criteria, offering a unique resource beyond diagnostic categories (Garg et al., 2023; Garg et al., 2024). BERT-based models trained on LoST achieved up to 85% F1 for sampled data with data augmentation, though performance declined in imbalanced settings, highlighting the subtlety of self-esteem cues.

Generating Synthetic Clinical Notes for Mental Health: Due to data scarcity and privacy concerns, synthetic clinical note generation has become a viable strategy for enabling NLP research in healthcare (Li et al., 2021). Early rule-based systems like SynthNotes focused on producing structurally realistic psychiatric notes, while recent advancements have leveraged LLMs and prompt engineering to generate more contextual text (Begoli et al., 2018). Models such as HEAL and GatorTronGPT have demonstrated that synthetic data can support downstream tasks like named entity recognition with performance comparable to models trained on real notes (Ive, 2022; Kang et al., 2024; Peng et al., 2023). Intrinsic and functional evaluations, including clinician reviews and BLEU scores, have shown that high-quality synthetic notes can closely mirror real clinical documentation (Li et al., 2021; Peng et al., 2023). A key measure of semantic fidelity is whether models trained on synthetic data perform comparably to those trained on real clinical notes. Past studies demonstrated that synthetic-trained models can rival real-trained models in tasks such as named entity recognition (Li et al., 2021; Peng et al., 2023). The existing synthetic corpora predominantly emphasizes clinical accuracy (e.g., diagnoses, medications), often lacking coverage of psychosocial constructs such as low self-esteem. To date, no synthetic datasets have been purposefully designed to construct them from user-generated inputs in synthetic data. To this end, a recent review emphasized the need for benchmarking synthetic text specifically on preserving mental health-related content (Ive, 2022). This highlights a critical gap and underscores the need for benchmarking synthetic data on its ability to preserve psychological and emotional depth in clinical contexts.

Social media to Clinical Contexts: Linking social media language to clinical outcomes, such as in Facebook-based studies predicting depression diagnoses, demonstrates the potential of social data for early mental health detection (Eichstaedt et al., 2018). However, direct model transfer between social media and clinical narratives remains challenging due to domain shift - differences in language style, tone, and narrative perspective. Social media posts are informal, first-person, and emotionally expressive, while clinical notes are formal, third person, and filtered through clinical observation. This mismatch can lead to semantic misalignment, where models trained on social data fail to recognize equivalent clinical expressions. Most domain adaptation work focuses on general biomedical vs. clinical text, with limited efforts bridging social and clinical domains (Laparra et al., 2021). Given the lack of labeled clinical corpora for constructs like low self-esteem, social media offers a promising proxy for synthetic health data generation. Still, the structural variation and semantic drift in social and clinical data for NLP is understudied.

Clinical Mental Health NLP: Clinical NLP methodologies in mental health have evolved from rule-based systems and keyword lexicons (Zhu et al., 2019) to advanced deep learning and transformer models (Rickman et al., 2025). While early approaches relied on pattern matching, recent studies demonstrate that transformer-based models, such as fine-tuned BERT and instruction-tuned LlaMA, significantly outperform traditional methods, particularly for detecting psychosocial factors like loneliness and anhedonia (Vance et al., 2025). Despite these advances, NLP has been limited to extract psychosocial constructs like social isolation and loneliness from EHRs, factors often missing from structured data (Myers et al., 2025; Rickman et al., 2025; Zhu et al., 2019). However, there is a significant lack of annotated datasets for interpersonal constructs such as low self-esteem in clinical notes. Privacy concerns and data-sharing restrictions further constrain clinical NLP research, prompting interest in synthetic note generation. This field lacks standardized benchmarks for underrepresented constructs like self-esteem or hopelessness, limiting comparative evaluation and clinical applicability (Chancellor & De Choudhury, 2020; Ive, 2022). Most available clinical NLP resources focus on tasks like entity recognition or diagnosis extraction rather than internal psychosocial states (Smith et al., 2020). This gap is largely due to the high cost and privacy concerns associated with creating expert-annotated clinical datasets.

Our study fills this gap by harnessing social media data to curate context-enhanced synthetic clinical notes, with a particular focus on psychosocial constructs such as low self-esteem. With this, we introduce a novel pipeline that leverages publicly available social media datasets to fine-tune language models for clinical applications and benchmark them against current state-of-the-art systems. This approach directly addresses the prohibitive costs and privacy risks inherent in manual chart reviews of clinical records. The innovation of our study lies in demonstrating how carefully engineered synthetic notes, derived from the linguistic and psychosocial richness of social media narratives, can approximate the contextual depth of clinical documentation.

## 2 Article type

In this section, first, we discuss study design. Second, we explain prompt engineering methods on small language models (Garg et al., 2025) for user-generated data to synthetic note generation pipeline, thereby transforming informal Reddit narratives into clinical-style notes. The generated synthetic notes were structured to ensure alignment with standard clinical documentation (Krishna et al., 2021; Ramprasad et al., 2023). Third, we present a stylometric analysis to characterize differences between Reddit posts and synthetic notes. Finally, we describe how to identify LoST in texts by deploying a parameter-efficient fine-tuning (Han et al., 2024) and assess its effectiveness in detecting low self-esteem across both Reddit posts and synthetic clinical notes.

### 2.1 Study design

For synthetic clinical note generation, we used the LoSTv2 dataset, a well-established resource curated from the subreddits r/depression and r/suicidewatch between December 2, 2021 and January 4, 2022 (Garg et al., 2024). The dataset comprises 2,174 user-authored posts, of which approximately 465 (∼25%) were manually annotated with textual cues reflecting LoST indicators [CITE LoSTv2]. For instance, ‘I am good for nothing’, ‘I can never become healthy again’. To mitigate this, we applied random oversampling to the minority class of classification labels within the training data only. This produced a balanced training set of 2,726 observations (50% positive, 50% negative). Oversampling was performed after the train/test split to avoid leakage, with a fixed random seed for reproducibility. Evaluation metrics were computed on testing dataset for identifying LoST. We used the full dataset for our experiments. These annotations provided contextual signals that were crucial for guiding our generation framework. We employed LLaMA 3.1 8B Instruct, a small instruction-tuned language model for synthetic note generation. Each user-authored Reddit post was treated as narrative input (*x*_*i*_), while the corresponding LoST annotation served as contextual information (*c*_*i*_). Together, these inputs were used to construct prompts designed to simulate physician-generated notes from given narratives. To control the truncation of synthetic notes, explicit length constraints were embedded within the prompt of our experiments. This structure ensured that outputs were faithful to patient-authored content, preserved psychosocial context, and adhered to the clinical documentation style. The reddit post is converted to synthetic notes (See Appendix A for example). During prompt engineering, we systematically compared different prompts to identify those yielding the most coherent and clinically plausible outputs. For comparison of synthetic notes with existing clinical documentation, we incorporated two additional resources: (i) the MediNote dataset (EPFL-IC-Make-Team/ClinicalNotes)1 (Kim et al., 2024), derived from 167K PubMed Central case reports and sampled initial synthetic dialogues (n=2,000 notes), and (ii) the clinical notes randomly selected from the Mayo Clinic Study of Aging (MCSA) cohort (n=1,160 notes) (Petersen et al., 2010), which is a population-based study cohort focused on understanding cognitive aging. To rigorously assess the fidelity and clinical utility of the generated synthetic notes, we designed a mixed-method evaluation strategy encompassing four dimensions. We conducted a stylometric analysis across four datasets to evaluate the linguistic characteristics of synthetic clinical text relative to authentic notes.

### 2.2. Context-enhanced Synthetic Health Data Generation

Our proposed method aims to generate context-enhanced synthetic health data by transforming user-generated narratives (e.g., Reddit posts expressing psychosocial concerns) into SOAP (Subjective, Objective, Assessment, Plan) format (Cameron & Turtle‐Song, 2002) where patient articulates concerns and physician documents them systematically. This framework is designed to preserve psychosocial context (e.g., low self-esteem cues) and enhance utility for downstream clinical NLP tasks while ensuring privacy. To enable context-aware generation, we represent each input narrative as a tuple of raw text, structured outcomes, and textual cues. Formally, let

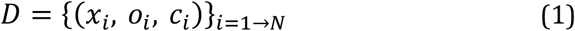

Where *x_i_* are a patient-authored narrative and *o_i_* refers to the associated outcomes (e.g., presence of low self-esteem as 1) and N are the number of instances. Equation 1 ensures that each narrative is paired with h outcome and the markers of contextual cues *c^i^* explicitly preserved and later infused into the generative process to produce clinically meaningful outputs.

#### Contextual representation

To ensure that synthetic health data captures not only surface-level content but also the underlying psychosocial nuance, we designed a multiple ‘components’ in contextual representation that integrates:

- Narrative Layer (*x_i_*)– This corresponds to the raw patient-authored narrative, which represents the subjective voice of the individual (e.g., a self-expressed concern drawn from online discourse).
- Psychosocial cues Layer (*c_i_*) – This layer encodes the contextual annotation (e.g., a low self-esteem marker or related psychosocial construct). Serving as an explicit signal, it anchors the generative process to clinically relevant cues that might otherwise be abstracted away.
- Structural Layer – To align generated text with clinical practice, we imposed constraints derived from the SOAP schema – Subjective, Objective, Assessment, and Plan.

These three components are integrated into a structured input prompt that guides LLAMA 3.1. 8B Instruct model to ensure that each generated note is conditioned simultaneously on the patient’s own words, the psychosocial context annotation, and the structural requirements of physician documentation. In doing so, it simulates the clinical process in which a patient expresses concerns, and the physician organizes them into standardized, structured notes.

#### Prompt engineering and model development

In line with the well-established literature, we deployed prompt engineering (Meskó, 2023). A small instruction-tuned language model is employed to generate structured clinical notes from user-generated narratives for reduced hallucinations and contextual assumptions. The model is framed as a conditional generator that maps an input narrative and its associated context into a SOAP-formatted note. Formally, for each sample we define:

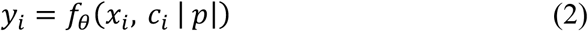

Where *yi* is the generated SOAP note and *f*(*θ*) denotes the language model parameterized by *θ* and prompt p. Equation 2 is employed to curate context-enhanced synthetic notes in response to the self-reported narratives. Both context and self-reported narratives are given as an input, supplemented by the engineered prompt p.

Our experiments indicate that the short, directive prompts yielded higher coherence and clinical fidelity than long, descriptive prompts. To address truncation, an explicit length of constraint must be incorporated during prompt construction, ensuring that outputs remain within a specified word limit. To minimize hallucinations, the model must be restricted from introducing unsupported assumptions, thereby aligning generation strictly with the provided narrative and context. The prompt is given as:

Prompt:

System instruction: In response to the patient’s notes and explanation, generate a short but structured clinical note in SOAP (Subjective, Objective, Assessment, Plan) format. Do not add, delete or update any new episodes or context. Make sure to include the explanation in clinical notes. Keep the length of clinical notes to 500 words.

User prompt: The patient’s notes is [NOTE] and the explanation is [CUES].

The prompt contains ‘system instructions’ for describing the task, and ‘user prompt’ for user query. The user prompt contains NOTE (*x_i_*) – indicating the Reddit posts, and CUES (*ci*) – the textual spans indicating low self-esteem in NOTE, if any.

### 2.3. Stylometric Analysis

To rigorously assess the fidelity and clinical utility of the generated synthetic notes, we designed a mixed-method evaluation strategy encompassing four dimensions.

#### Structural Analysis

To understand the inherent differences between Reddit posts and synthetic notes, we conducted a structural analysis based on network representations of language. Two complementary networks were constructed for each dataset: (i) a word co-occurrence network (WCN), where nodes represent all unique words and edges denote co-occurrence within a context window, and (ii) an entity co-occurrence network (ECN), restricted to named entities (e.g., symptoms, conditions, psychosocial terms) and their co-occurrence obtained through NLTK and spacy libraries of Python. By comparing these networks, we characterized the global topology of each dataset, revealing how words and concepts are structurally connected. This analysis provided insights into differences in stylometry and narrative style across text types.

The basic measures are (i) the number of nodes - unique words or entities; (ii) the number of edges - co-occurrence links between them; and (iii) node-to-edge ratio further captures network sparsity or compactness, with lower values indicating tightly interconnected language, as is typical in clinical documentation, and higher values reflecting the looser associations common in informal Reddit text. In addition, we analyzed the overall connectivity and density of the network through seven network metrics. The average degree explains contextual interconnectedness; higher values can reflect repetitive structures often found in structured clinical notes. The clustering coefficient quantifies the extent to which nodes form tightly knit groups. Elevated clustering typically signals recurring phrase patterns or domain-specific jargon, while lower clustering points to more variable, less constrained language use. Network density, defined as the proportion of observed edges relative to all possible connections, distinguishes between highly repetitive discourse and more diverse narrative expression. The number of connected components indicates whether the network forms a single cohesive structure or multiple fragmented subgraphs. Degree assortativity measures whether nodes with many connections tend to link to one another. In natural language, this metric often takes a negative value, as highly frequent words link to rarer ones; however, the magnitude of this effect may vary between structured clinical writing and informal social media narratives. Finally, we assessed community structure using greedy modularity optimization. The number and size of detected communities reflect thematic organization within the text. A greater number of smaller communities suggest broad thematic diversity while fewer and larger communities are indicative of the more constrained discourse.

By systematically analyzing the topology of word and entity co-occurrence networks, we gain an objective understanding of how narrative style, lexical choices, and contextual connections vary across datasets. This structural lens offers a quantitative foundation for evaluating whether synthetic notes shift toward the stylometry of real-world clinical narratives.

#### Readability Analysis

To evaluate linguistic variation across datasets, we employed a set of established readability indices that capture different dimensions of text complexity. These measures are widely used in computational linguistics and education research to approximate the grade level, readability, and stylistic difficulty of written text. The Automated Readability Index (ARI) estimates text complexity based on characters per word and words per sentence, producing a score that corresponds to U.S. grade levels (Smith & Senter, 1967). Similarly, the Coleman–Liau Index (CLI) uses character counts rather than syllables, offering a computationally efficient readability estimate aligned to educational grade levels (Severance & Cohen, 2015). The complementary Flesch Reading Ease (FRE) score rates text on a scale from 0 to 100, with higher scores indicating easier readability (Massie et al., 2024). Finally, the Gunning Fog Index (GFI) estimates the number of years of formal education required to comprehend a text (Gross & Sadowski, 1985). Together, these indices allow for a nuanced assessment of linguistic complexity and stylistic variation between Reddit posts, synthetic notes, and clinical notes. Informal Reddit narratives are expected to score as more readable (higher FRE, lower grade-level indices), while clinical notes, with their dense medical terminology, typically yield higher grade-level scores and a higher Gunning Fog Index. Synthetic notes are anticipated to occupy an intermediate position, balancing accessibility with clinical formality.

#### Diversity

Lexical diversity was quantified using the type–token ratio (TTR), which measures the proportion of unique words to the total number of words across the corpus (Ratner et al., 2024). A higher TTR indicates a richer and more varied vocabulary, whereas lower values reflect greater repetition of the same lexical items. While simple and interpretable, this measure is sensitive to text length and may underestimate diversity in longer documents. To complement this surface-level analysis, we assessed semantic variability using embedding diversity. Here, we employed the Sentence-BERT model (Reimers & Gurevych, 2019) to generate dense vector representations of each text instance and computed pairwise cosine similarities between embeddings. The mean similarity was then inverted to produce a cosine dissimilarity score, such that higher values reflect greater semantic heterogeneity across the dataset. Finally, we evaluated overlap-based diversity using the Jaccard index, which compares the proportion of shared versus unique tokens (words) between pairs of texts. We report one minus the average Jaccard similarity, thereby capturing the extent to which texts diverge in their lexical content. Together, these three complementary metrics—lexical diversity, embedding diversity, and Jaccard-based diversity—provide a comprehensive assessment of both surface-level vocabulary variation and deeper semantic distinctions in the text corpus.

#### Contextual Fidelity

We implemented a small language model as a judge (SLM–as–a–judge) to assess whether textual cues listed in LoST are expressed in paired clinical notes (Psych_notes_LLAMA31) (J. Gu et al., 2024). The judge was Meta-Llama-3.1-8B-Instruct, run locally via Hugging Face Transformers with deterministic decoding (*max*_*new*_*tokens* = 512) and direct PyTorch generation. For each row, LoST strings of the form “text1, text2, text3” were normalized into a deduplicated cue list (split on commas/semicolons/pipes/newlines; whitespace-collapsed; case-insensitive comparison) where all text1, text2 and text3 are different LoST cues. Notes were head–tail clipped to a fixed character budget to control context length. The system instruction required a single JSON object with fields found_cues, missing_cues, and coverage_rate, and specified evidence-based criteria: a cue is “present” only if supported by a short verbatim span or an unambiguous paraphrase in context; explicit negation (e.g., “denies guilt”) is treated as absent. Coverage rate is the proportion of textual cues indicating LoST that are successfully identified or represented by the SLM-as-a-judge, relative to the total cues that should have been covered. The coverage rate is NaN if there are no textual cues indicating LoST in synthetic notes.

If coverage_rate was non-numeric, it was ignored and recomputed deterministically as 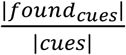. To prevent synonym drift, found_cues were snapped back to the original cue vocabulary (case-insensitive). The resulting fields (found_cues, missing_cues, coverage_rate) were appended to the dataframe for downstream analysis.

### 2.4. Binary classification of psychosocial symptoms

In this study, we fine-tuned a SLM, using a parameter-efficient adaptation strategy tailored for binary classification of LoST narratives (Dubey et al., 2024; J. Zhang et al., 2024). Rather than updating the full model, we keep the pretrained backbone frozen and introduce lightweight LoRA adapters into a targeted subset of transformer layers, coupled with a minimal classification head.

#### Base Model

The pretrained small language model used in this study is LLaMA-3.1-8B Instruct, a decoder-only transformer with approximately 8 billion parameters. This model belongs to the LLaMA (Large Language Model Meta AI) family (Vavekanand & Sam, 2024), which adopts a GPT-style autoregressive architecture trained with an objective to classify given text into the presence or absence of low self-esteem. We initialized our experiments using the publicly released Hugging Face checkpoint of LLaMA-3.1-8B Instruct.

Since LoST cues are often implicit and context-dependent, careful prompt engineering was critical in adapting LLaMA-3.1-8B Instruct for binary LoST classification. We adopted an instruction-tuning framework, where each training example was reformulated into a system–user–assistant dialogue template, consistent with the model’s pretraining style.

- System Instruction (SI): The system message establishes the role of the model as a clinical text classifier and specifies the rules of annotation. This high-level guidance ensures that the model consistently interprets task requirements across diverse training samples.
- User Prompt (*Pi*): The user message contains the clinical note or Reddit posts to be classified. To avoid label leakage, the prompt did not include ground-truth labels— these were only used during supervised training for loss computation.
- Assistant Response (*yi*): The assistant’s output is the gold-standard label, either presence (1) or absence (0) of LoST. This forms the supervised signal for fine-tuning.

The working instance of Reddit post for identifying low self-esteem is as follows:

System Instruction: You are an expert in identifying episodes of low self-esteem from a given text. You are given a patient’s note. Do not assume anything. Based only on the text, decide if the patient shows signs of LOW SELF-ESTEEM (1 = yes, 0 = no). Respond only in this format (1 or 0). Do not add any explanation. The note given is:

User prompt: I feel worthless most of the time and believe I am a burden to my family.

Assistant Response: 1

Formally, each training instance was structured as:

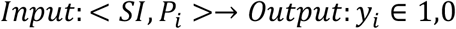

During fine-tuning, the system and user segments were concatenated into the model’s input sequence, while the assistant label was treated as the supervised target. By maintaining a consistent prompt template, we aligned our fine-tuning setup with the model’s original instruction-following behavior, thereby reducing training instability and enhancing generalization.

LoST Identification. For LoST identification, we employed low rank adaptation methods. LoRA achieves a balance by training only 0.1–1% of parameters without additional inference cost (Hu et al., 2022). LoRA is a parameter-efficient fine-tuning method that injects trainable low-rank decomposition matrices into transformer models while freezing all pre-trained weights. By doing so, the number of trainable parameters for downstream tasks is reduced from billions to a small fraction, enabling efficient adaptation without sacrificing performance. Let the pre-trained weight matrix be *W*0 = ℝ^*d*⋅*k*^ where d and k are the number of rows and columns of the matrix whose entries are real numbers. In the standard fine-tuning, one would update the full matrix *W*_0_. Instead, LoRA reduces the number of trainable parameters from (*d* ⋅ *k*) to *r*(*d* + *k*), and constrains the weight update Δ*W* = *BA* to a low rank decomposition where *A* ∈ ℝ^*r*⋅*k*^ and *B* ∈ ℝ^*d*⋅*r*^, and *r* ≪ min(*d, k*). Here, matrix A compresses the input (*k* → *r*) into a small subspace. and B expands it back (*r*→*d*) to match the original output size. The forward propagation then becomes

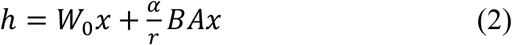

where *α* is the scaling hyperparameter and r is the rank. LoRA relies on a set of hyperparameters that jointly balance efficiency, stability, and adaptability. The rank controls the trade-off between expressivity and parameter efficiency, with higher ranks enabling richer task-specific representations at the cost of more trainable parameters. The scaling factor ensures that low-rank updates remain balanced and do not overwhelm the contribution of the frozen pre-trained weights, thereby stabilizing optimization. LoRA is applied selectively to the attention projection layers of language models, which are critical for capturing contextual dependencies. To prevent overfitting, especially on smaller datasets, LoRA dropout introduces stochastic regularization to the update pathways. The bias setting typically remains “none,” keeping bias parameters frozen to preserve the pre-trained distribution and reduce training complexity. The model is optimized using the standard cross-entropy loss, which maximizes the likelihood of the correct target tokens given input prompts. This objective aligns the low-rank updates with task-specific supervision while preserving the knowledge encoded in the frozen base weights.

## 3 Results

To assess the fidelity and utility of the generated notes, we conducted a mixed-method evaluation strategy comprising four complementary dimensions: (i) structural analysis, (ii) readability analysis, (iii) linguistic diversity, and (iv) contextual fidelity of LoST cues between the source Reddit posts and the corresponding synthetic notes. We also evaluated our approach for detecting LoST in synthetic clinical narratives and Reddit posts using the fine-tuned LLaMA-3.1-8B Instruct model with LoRA adapters.

### 3.1 Synthetic Health data generation

#### 3.1.1 Structural Analysis

A comparison of network metrics across Reddit (LoSTv2), Synthetic Health Data Generated (SHDG), MediNote, and Mayo Clinic notes highlight several key observations as shown in Table 1.

**Table 1.** Structural variations among different datasets through network metrics.

| Metric | Reddit |  | SHDG |  | MediNote |  | Mayo Notes |  |
| --- | --- | --- | --- | --- | --- | --- | --- | --- |
|  | ECN | WCN | ECN | WCN | ECN | WCN | ECN | WCN |
| Nodes | 1,904 | 8,842 | 1,743 | 8,590 | 16,223 | 25,849 | 3,925 | 6,897 |
| Edges | 3,405 | 65,109 | 4,360 | 104,223 | 45,560 | 316,262 | 8,560 | 35,019 |
| Node:Edge Ratio | 0.56 | 0.14 | 0.40 | 0.08 | 0.36 | 0.08 | 0.46 | 0.20 |
| Average Degree | 3.58 | 14.73 | 5.00 | 24.27 | 5.62 | 24.47 | 4.36 | 10.15 |
| Clustering Coefficient | 0.05 | 0.19 | 0.28 | 0.26 | 0.10 | 0.21 | 0.13 | 0.10 |
| Density | 0.0019 | 0.0017 | 0.0029 | 0.0028 | 0.0003 | 0.0009 | 0.0011 | 0.0015 |
| Connected Components | 44 | 1 | 9 | 1 | 1 | 1 | 4 | 1 |
| Degree Assortativity | -0.05 | -0.13 | -0.24 | -0.11 | -0.09 | -0.10 | -0.09 | -0.05 |
| Communities (Greedy) | 79 | 183 | 41 | 164 | 287 | 395 | 103 | 107 |

In general, the number of nodes and edges are fewer for ECN as compared to WCN due to selective entities in ECN. The node-to-edge ratio was higher in ECNs than in WCNs, reflecting that entity-level networks capture meaningful, relevant connections, whereas word-level networks are influenced by randomized lexical co-occurrences. Reddit shows the highest ratio for ECN (0.56) whereas the ratio of SHDG is closer to MediNote and Mayo Notes suggesting that synthetic notes structurally approximate clinical narratives more closely than Reddit posts.

SHDG has the highest clustering coefficient in entity networks (0.28), suggesting more tightly connected entity clusters, possibly reflecting repetitive phrasing during generation. Clinical datasets have a higher clustering coefficient than Reddit but much lower than SHDG. SHDG shows the high density whereas density of clinical datasets is markedly lower, reflecting the breadth and sparsity of large, detailed clinical case reports. This implies that synthetic notes may introduce over-regularization of connections, possibly due to language model tendencies toward repetitive co-occurrence. Similar observations are made for assortativity. Reddit generally is highly fragmented as compared to SHDG and other clinical datasets, reflecting diverse, unstructured discourse. This shows synthetic generation increases cohesion and standardization, reducing narrative fragmentation. Clinical notes contain large numbers of communities as compared to Reddit and SHDG, reflecting the thematic richness of real clinical notes. Our findings indicate that, at a structural level, synthetic notes exhibit greater similarity to authentic clinical narratives, whereas Reddit-derived data remains more distinct in its stylometric profile. Nonetheless, further investigation is warranted to address discrepancies in density and clustering coefficients, potentially through strategies that enhance data diversity.

#### 3.1.2 Readability Analysis

The readability analyses highlight how closely synthetic notes align with authentic clinical documentation, as opposed to the informal style of Reddit posts in terms of readability (Table 2).

**Table 2.** Readability analysis among different datasets.

| Metric | Reddit<br>(2,173) | SHDG<br>(2,173) | MediNote<br>(2,000) | Mayo Notes<br>(1,160) |
| --- | --- | --- | --- | --- |
| Automated Readability Index | 7.27 | 15.13 | 13.70 | 14.87 |
| Coleman–Liau Index | 5.44 | 14.60 | 14.01 | 13.76 |
| Flesch Reading Ease | 75.08 | 33.04 | 34.23 | 30.00 |
| Gunning Fog Index | 9.35 | 16.49 | 16.30 | 16.87 |

On the ARI, Reddit posts scored 7.27 indicating a middle school level of accessibility. In contrast, SHDG (15.13), MediNote (13.70), and Mayo Notes (14.88) all reflected university-level difficulty, highlighting a distinct shift from informal to professional language. A similar pattern was observed with the CLI. These results suggest that synthetic notes align more closely with authentic clinical datasets than with Reddit in terms of lexical density. The GFI showed Reddit narratives at 9.35, indicating high school readability, compared to SHDG (16.49), MediNote (16.30), and Mayo Notes (16.87), which are aligned with professional or academic-level texts. Across all indices, the clinical datasets and SHDG clustered tightly, while Reddit stood apart as substantially more accessible.

#### 3.1.3 Diversity

Three diversity metrics highlight notable distinctions of linguistic diversity between Reddit, synthetic clinical note, and real clinical notes (MediNote and Mayo notes) as shown in Table 3.

**Table 3.** Diversity among different datasets.

|  | Reddit | SHDG | MediNote | Mayo notes |
| --- | --- | --- | --- | --- |
| *Lexical Diversity (Type-Token Ratio)* | 0.07 | 0.03 | 0.06 | 0.08 |
| *Jaccard-Based Diversity* | 0.91 | 0.85 | 0.91 | 0.96 |
| *Embedding Diversity (Cosine Dissimilarity)* | 0.71 | 0.46 | 0.68 | 0.70 |

Lexical diversity, as measured by the type–token ratio, is higher for Mayo notes (0.0778), MediNote (0.0628) and Reddit posts (0.0717), indicating richer vocabulary usage relative to the Synthetic notes (0.0275), which exhibit substantial repetition of terms. The higher Jaccard-based diversity scores for Mayo notes, MediNote and Reddit data as compared to Synthetic notes further underscore this contrast. This indicates that synthetic notes may rely more heavily on standardized clinical expressions, which constrains their overlap-based variability. Finally, embedding diversity, which captures semantic variety beyond surface-level word use, reveals greater heterogeneity in Reddit posts (0.7184), Mayo notes (0.6997) and MediNote (0.6808). Synthetic notes again exhibit the lowest score (0.4644), reflecting their limited semantic variation. As such, we noticed comparatively lower lexical and semantic diversity of synthetic data as that of Reddit and existing clinical notes. Models trained on this homogeneous data may be learning repetitive output of the generator LLM. We keep improvements in this area are the future research direction.

#### 3.1.4 Contextual Fidelity

To quantify the contextual enhancement of synthetic notes, we measure the contextual coverage of textual cues indicating LoST in synthetic notes. The coverage distribution indicates high overall fidelity (mean ≈ 0.81), with a pronounced ceiling effect: the 75th percentile at 1.0 implies at least a quarter of instances achieve perfect coverage. The central spread is moderate (IQR = 0.4), spanning from Q1 = 0.6 to Q3 = 1.0, meaning the middle half of items ranges from “good” to “perfect.” This pattern suggests a right-censored distribution with mass at the upper bound and a lower tail at or below 0.6, pointing to residual heterogeneity across cases. Figure 2 demonstrates the cumulative distribution of coverage rate.

**Figure 2.**
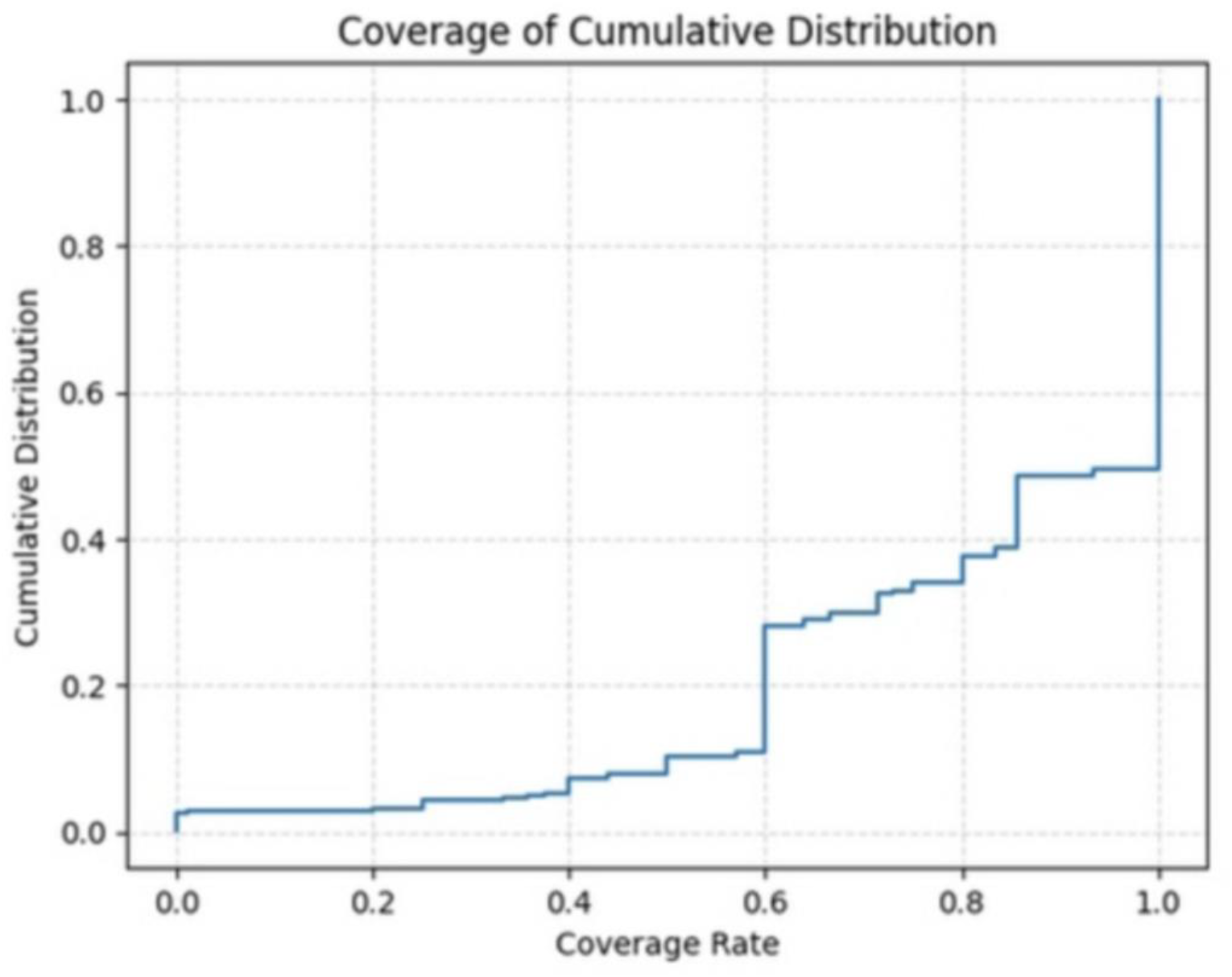
Coverage of cumulative distribution

We noticed that synthetic notes are generally successful in preserving contextual information, yet the presence of a lower tail suggests inconsistency in coverage rate. Approximately 51% of the instances achieved full coverage in contextual enhancement, ∼20% lies between 60% to 80% coverage, while another 18% of the remaining cases exhibited a coverage rate of around 60%. Few synthetic notes (∼11%) may contribute disproportionately to the lower coverage scores (<60%). This highlights that while the generation pipeline performs strongly in aggregate, further refinement may be needed to better handle sparse or ambiguous inputs.

### 3.2 Experimental results - Identifying low self-esteem (LoST)

Across both datasets, as shown in Table 4, LoRA fine-tuning delivers the largest gains on all summary metrics, with especially strong improvements on SHDG.

**Table 4.**
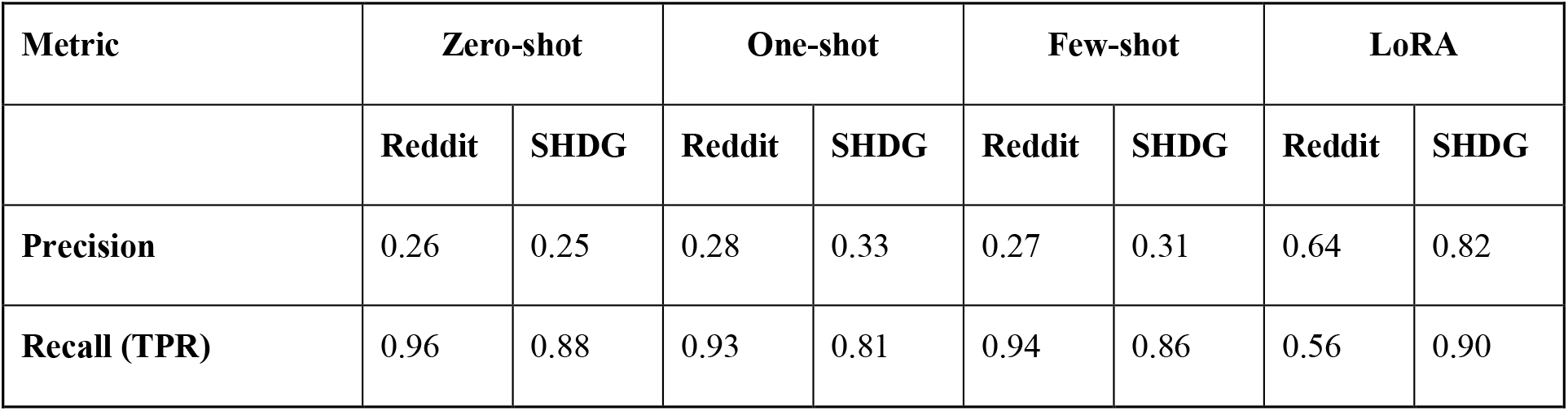
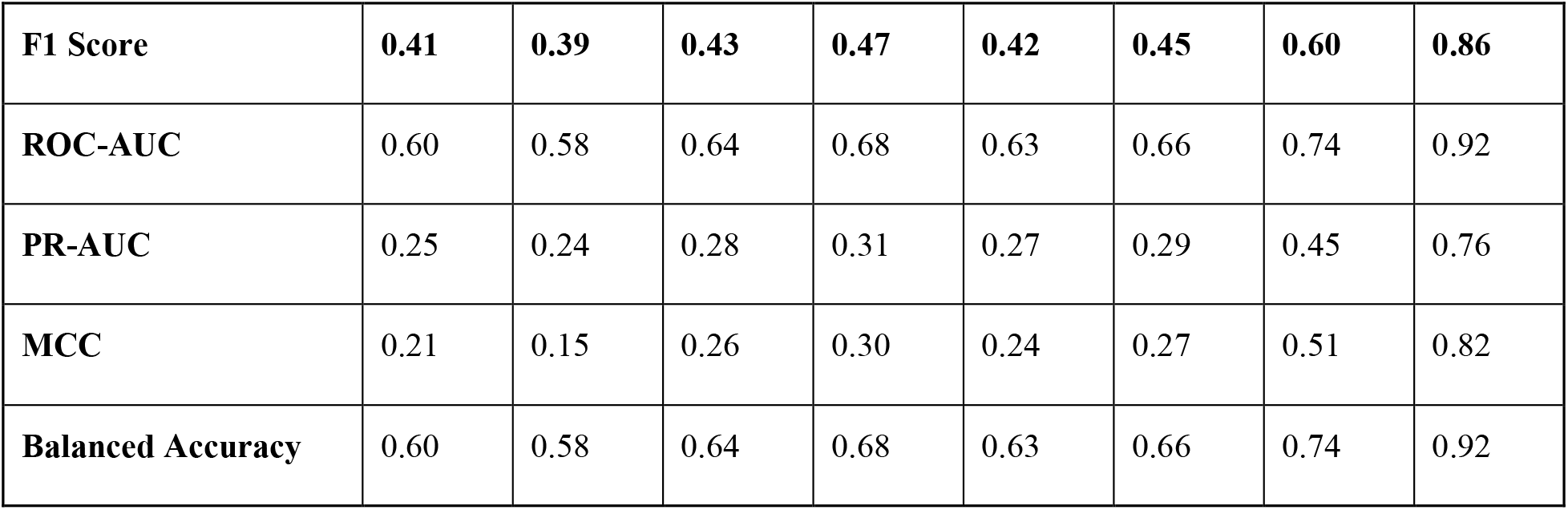
Experimental results for identifying LoST in Reddit posts and synthetic notes.

| Metric | Zero-shot |  | One-shot |  | Few-shot |  | LoRA |  |
| --- | --- | --- | --- | --- | --- | --- | --- | --- |
|  | Reddit | SHDG | Reddit | SHDG | Reddit | SHDG | Reddit | SHDG |
| Precision | 0.26 | 0.25 | 0.28 | 0.33 | 0.27 | 0.31 | 0.64 | 0.82 |
| Recall (TPR) | 0.96 | 0.88 | 0.93 | 0.81 | 0.94 | 0.86 | 0.56 | 0.90 |
| **F1 Score** | **0.41** | **0.39** | **0.43** | **0.47** | **0.42** | **0.45** | **0.60** | **0.86** |
| **ROC-AUC** | 0.60 | 0.58 | 0.64 | 0.68 | 0.63 | 0.66 | 0.74 | 0.92 |
| **PR-AUC** | 0.25 | 0.24 | 0.28 | 0.31 | 0.27 | 0.29 | 0.45 | 0.76 |
| **MCC** | 0.21 | 0.15 | 0.26 | 0.30 | 0.24 | 0.27 | 0.51 | 0.82 |
| **Balanced Accuracy** | 0.60 | 0.58 | 0.64 | 0.68 | 0.63 | 0.66 | 0.74 | 0.92 |

Prompting alone (zero/one/few-shot) yields modest differences (around 2% to 4% of F1-score) on the Reddit data, but one- and few-short learning improved the performance against zero-short learning in SHDG. In SHDG, LoRA improved the performance a lot, suggesting that SHDG is well aligned with fine-tuning rather than in-context hints alone. In Reddit data, LoRA made the model substantially stricter—precision rises, but recall falls, but overall performance improved (higher F1 and AUCs). As shown in Figure 3, the training curves for both datasets show steady optimization; loss decreases continuously from the earliest to the final recorded steps.

**Figure 3.**
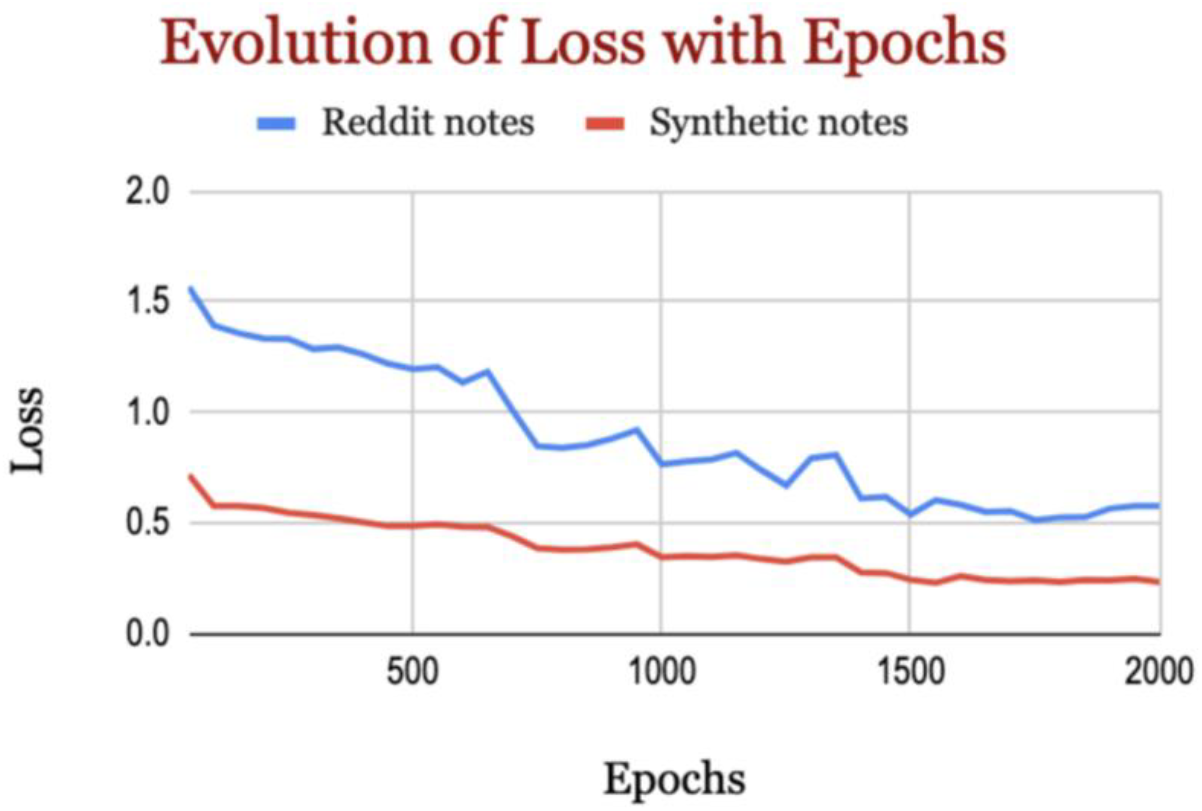
Evolution of loss with epochs.

On Reddit, training loss falls from about 1.568 at the beginning to roughly 0.576 at the end, a reduction of about 63%. On synthetic data, loss starts far lower—around 0.717—and descends to about 0.233, a reduction of about 67%, substantially lower loss than the Reddit model, indicating an easier objective and/or cleaner supervision in that domain.

## 4 DISCUSSION

Reddit posts provide raw, informal expressions of psychosocial constructs that are often under-documented in clinical notes. However, their informal style, fragmented structure, and lack of clinical framing limit their direct applicability for training clinical NLP systems. By generating synthetic notes from Reddit narratives, we simulate how such patient experiences would be represented in a clinical environment. Our model acts as a “bridge,” mapping between raw patient expression and the structured, clinician-oriented language of EHRs. This simulation is significant for two reasons. First, synthetic notes mitigate privacy concerns inherent in real patient records, enabling researchers to create large-scale, shareable datasets without exposing protected health information. Second, contextually enhanced synthetic notes provide training material that is closer in format to actual EHRs, making them practically useful for clinical NLP model development.

The applicability of synthetic notes extends across multiple domains of clinical NLP. They provide training and evaluation resources for tasks such as concept extraction, where annotated data is limited or costly to obtain. During prompt engineering for synthetic notes generation, we observed that concise, directive prompts generated higher-quality notes than verbose, heavily contextualized ones. Second, synthetic notes retain structural and stylistic similarities to real documentation while allowing controlled variability in contextual diversity.

Our findings confirm that Reddit data is structurally distinct than synthetic notes which demonstrate structural metrics that are closer to MediNote and Mayo clinical notes. However, SHDG also shows higher clustering and density, which may indicate artifacts of language model generation such as repetitive phrases and reduced diversity that differentiate them from authentic clinical documentation. However, further clarification is needed regarding the cause behind observed reduction in diversity of language model outputs, and whether this effect is specifically associated with certain model architectures, such as LLaMA. Reddit narratives are consistently simpler, more readable, and conversational, reflecting informal social discourse. By contrast, SHDG, MediNote, and Mayo note clusters tightly across all indices, indicating greater linguistic complexity and technicality, characteristic of professional clinical documentation. Importantly, SHDG shows structural and readability alignment with clinical notes, supporting its utility as a proxy training resource while also confirming its departure from the informal stylometry of Reddit data.

Clinical notes vary considerably in their lexical and semantic richness, with Mayo notes showing unexpectedly high diversity across multiple dimensions. In contrast, synthetic notes are the most homogeneous, reflecting the templated and repetitive nature of generated documentation. These distinctions have important implications for clinical NLP pipelines, as models trained on inappropriately constructed synthetic notes may fail to generalize to more linguistically and semantically diverse corpora such as Mayo notes. The observed contextual variation across segments highlights the importance of accounting for lexical diversity. This underscores the need for more robust synthetic note generation methods that explicitly incorporate these features. Our observations indicate a clear domain shift in linguistic complexity from Reddit to synthetic notes. Longer sentences, denser terminology, and medical vocabulary make both synthetic and clinical notes substantially harder to read than lay discourse. For NLP systems moving across these sources, domain adaptation is advisable, including acronym expansion, vocabulary normalization, sentence segmentation, and tuning tokenization to clinical stylometry. Our synthetic data generation method demonstrates strong alignment with real-world data in terms of contextual fidelity, structural coherence, and readability. Nonetheless, future work should focus on enhancing diversity to further improve the overall quality of the data generated to fully support generalizability.

The base model – (LLAMA 3.1 8B Instruct) is operated in 16-bit floating point precision to ensure numerical stability and prevent degradation from low-bit arithmetic. By restricting LoRA to the last few layers, the model adapts just enough to capture subtle self-esteem cues in text while keeping the trainable parameter count small. Moving from zero- to one-shot generally helps a bit; moving to few-shot does not consistently outperform one-shot. Improvements with LoRA are not confined to a single operating point: precision–recall curves shift favorably, and overall separability improves. Thus, we demonstrated the effectiveness of LoRA fine-tuning for identifying LoST in synthetic notes and recommend tuning language models for identifying psychosocial concepts in text.

In Reddit data, LoRA outperformed the n-shot learning models in identifying LoST, at the expense of recall. This pattern is typical when a noisy, heterogeneous domain is regularized by fine-tuning: the model stops over-flagging but may miss some positives at the default threshold. Post-hoc threshold tuning on a Reddit-like validation set is advisable to recover recall while keeping the precision gains. Re-tuning the decision threshold (or use cost-sensitive tuning) could be applied to trade some precision for higher recall without losing LoRA’s robustness. If Reddit recall remains critical, domain-targeted augmentation or a small Reddit-specific LoRA may be considered to reduce domain shift.

This study has several limitations. First, the annotated real clinical notes are not yet available for evaluation, limiting the assessment of LoST classification in clinical settings. Second, while the generated synthetic clinical notes follow a SOAP structure, the “Objective” section often reflects subjective information, and our current model has limited capability to accurately represent the boundaries between them.

Our study demonstrated the use of publicly available mental health datasets to curate synthetic clinical notes that can be leveraged for training NLP models on downstream tasks such as entity recognition and concept extraction. This approach could reduce reliance on manual chart review by clinicians and medical experts, thereby improving resource utilization in NLP development. While the contextually enhanced synthetic notes exhibit high stylometric fidelity with real clinical documentation, they remain limited by low diversity in the source of data, which constrains the quality of data curation. High-quality data curation and integration into language model tuning hold significant potential for improving model development and performance.

## Author Contributions

MG: Conceptualization, Data curation, Formal analysis, Methodology, Writing – original draft. XL, EJ: Validation, Writing – review & editing. JB, MF: Investigation, Project administration, Writing – review & editing. YG: Validation, Writing – review & editing. SS: Conceptualization, Investigation, Project administration, Resources, Funding acquisition, Writing – review & editing, Visualization.

## Generative AI statement

The author(s) declare that no Generative AI was used in the creation of this manuscript.

## Data Availability Statement

The publicly available Reddit dataset which is analyzed for this study can be found at http://github.com/drmuskangarg/LoST.

